# The Great Recanalization Debate in Acute Ischemic Stroke: Direct Thrombectomy versus Bridging Therapy—A Meta-analysis of Randomized Controlled Trials

**DOI:** 10.64898/2026.05.10.26352784

**Authors:** Adila Jawaid, Manabesh Nath, Shubham Misra, Deepti Vibha, Pradeep Kumar

## Abstract

**Background:** Endovascular thrombectomy (EVT) is the standard of care for acute ischemic stroke caused by large-vessel occlusion. However, the additional benefit of intravenous thrombolysis (IVT) before EVT remains controversial. This systematic review and meta-analysis evaluated the efficacy and safety of bridging therapy (EVT plus IVT) compared with EVT alone.

**Methods:** This systematic review and meta-analysis was conducted according to PRISMA 2020 and Cochrane Handbook recommendations and prospectively registered in PROSPERO. PubMed, EMbase, Scopus, and the Cochrane Library were searched for randomized controlled trials published between 1^st^ January 2015 and 30^th^ April 2026 comparing EVT plus IVT versus EVT alone in acute ischemic stroke. Random-effects meta-analysis was performed to estimate pooled odds ratios (ORs) with 95% confidence intervals (CIs). Primary outcomes included functional independence at 90 days and successful recanalization. Secondary outcomes included symptomatic intracranial hemorrhage (sICH) and all-cause mortality.

**Results:** Eleven randomized controlled trials involving 4,419 patients were included in the meta-analysis. Compared with EVT alone, bridging therapy was associated with significantly better functional independence at 90 days (OR=1.25; 95% CI: 1.02–1.53). Patients receiving EVT plus IVT also demonstrated a trend toward higher rates of successful recanalization (OR=1.25; 95% CI: 0.95–1.64) and lower 90-day mortality (OR=0.84; 95% CI: 0.67–1.04). The risk of sICH was comparable between the two treatment strategies (OR=1.07; 95% CI: 0.81–1.40). Overall, the certainty of evidence was rated as moderate.

**Conclusions:** Bridging therapy before EVT may improve functional outcomes and recanalization without increasing sICH, supporting its use as a reasonable treatment strategy in eligible patients with acute ischemic stroke.

## Introduction

Acute ischemic stroke (AIS) due to large vessel occlusion (LVO) represents a distinct clinical and pathophysiological entity associated with high rates of mortality and long-term disability [1]. LVO strokes typically result in abrupt cessation of blood flow to large cortical territories, leading to rapid infarct core expansion, impaired collateral circulation, and early neurological deterioration [2,3]. Therefore, timely and durable recanalization is the most critical determinant of functional outcome in this patient population [4].

Endovascular thrombectomy (EVT) has fundamentally altered the management of AIS due to LVO by enabling rapid mechanical recanalization of occluded intracranial arteries [5]. Randomized controlled trials (RCTs) have consistently demonstrated that EVT improves rates of successful recanalization and functional independence when performed within the appropriate window period and in accordance with patient eligibility [6–10]. Nevertheless, mechanical recanalization does not uniformly translate into optimal tissue recanalization, as microvascular obstruction, distal embolization, and no-reflow phenomena may limit clinical benefit [11].

Intravenous thrombolysis (IVT), administered before EVT—referred to as bridging therapy—has been implemented to address these limitations. From a biological standpoint, IVT may promote early partial recanalization, reduce thrombus burden, alter clot composition, and improve distal microcirculatory flow, potentially improving the effectiveness of subsequent thrombectomy [12]. However, IVT may also increase the risk of hemorrhage transformation, particularly in the setting of large infarct cores or delayed recanalization, and may cause thrombus fragmentation with distal embolization [13]. In addition, concerns have been raised regarding treatment delays and resource utilization, especially in high-volume comprehensive stroke centres where EVT can be initiated expeditiously [14].

Previous meta-analyses[15–21] comparing bridging therapy (EVT + IVT) versus EVT alone have shown heterogeneous results, reflecting variations in thrombectomy devices, thrombolytic agents and dosages (e.g., alteplase vs. tenecteplase), neuroimaging selection, onset-to-treatment times, and definitions of clinical and safety outcomes. This variability has contributed to inconsistent clinical practice and uncertainty in guideline recommendations, highlighting the need for a rigorous synthesis of high-quality evidence. To address this clinical equipoise, we conducted a systematic evaluation of published RCTs comparing bridging therapy with EVT alone in patients with acute ischemic stroke due to large vessel occlusion, aiming to clarify the balance of efficacy and safety across clinically meaningful outcomes and inform evidence-based decision-making.

## Materials and Methods

### Study design and protocol

This systematic review and meta-analysis of RCTs was conducted to compare EVT with and without prior thrombolysis in patients with acute ischemic stroke due to large vessel occlusion. The review was performed in accordance with the PRISMA 2020 [22] reporting guidelines and the Cochrane Handbook for Systematic Reviews of Interventions [23]. The protocol was prospectively registered in the PROSPERO database (Registration ID: **CRD420251042070**) to ensure methodological transparency and reduce the risk of selective reporting.

### Data sources and search strategy

A comprehensive literature search was performed in PubMed, EMBASE, Scopus, and the Cochrane Central Register of Controlled Trials (CENTRAL) to identify eligible studies published between 01^st^ January 2015 and 30^th^ April 2026. The search strategy combined Medical Subject Headings (MeSH) and free-text terms related to acute ischemic stroke, large vessel occlusion, endovascular therapy, mechanical thrombectomy, bridging therapy, intravenous thrombolysis, and randomized controlled trials. The complete search strategy is provided in the Supplementary Material (**Supplementary Table 1**). In addition, reference lists of included articles and relevant reviews were manually screened, and abstracts from major international conferences, including the International Stroke Conference and the European Stroke Organisation Conference, were reviewed to identify unpublished or ongoing trials.

### Eligibility criteria

Studies were included if they met the following criteria: (1) enrolled adults (≥18 years) with acute ischemic stroke attributable to large vessel occlusion involving the intracranial internal carotid artery, middle cerebral artery (M1 or M2 segments), or basilar artery; (2) compared bridging therapy EVT + IVT v*s.* EVT alone; (3) reported at least one predefined efficacy or safety outcome; and (4) used a randomized controlled trial design. Studies were excluded if they were non-randomized, observational in nature, case reports or case series, involved paediatric populations, or did not provide sufficient data for effect estimation.

### Outcomes

The primary efficacy outcomes were functional independence, defined as a modified Rankin Scale (mRS) score of 0–2 at 90 days, and successful recanalization, described as a Thrombolysis in Cerebral Infarction (TICI) score of 2b–3. Safety outcomes included symptomatic intracranial haemorrhage (sICH) within 24 hours and all-cause mortality at 90 days.

### Data extraction

Two reviewers (AJ and MN) independently screened titles and abstracts, assessed full-text articles for eligibility, and extracted data using a standardized data extraction form. The extracted data included study characteristics, baseline patient demographics, treatment protocols, baseline investigations and clinical assessments, radio-imaging criteria, treatment measures, and detailed post-intervention and long-term follow-up outcomes. Disagreements were resolved through discussion and consensus, with adjudication by the corresponding author (PK) when required.

### Risk of bias

Risk of bias for individual included RCTs were assessed using the Cochrane Risk of Bias Tool version 2.0 [24]. which assesses five domains: bias arising from the randomization process, deviations from intended interventions, missing outcome data, measurement of outcomes, and selection of reported results. Each domain was rated as low risk, some concerns, or high risk, and an overall judgment was assigned to each trial.

### Certainty of Evidence

The certainty of evidence for each outcome was assessed using the GRADE approach and summarized with GRADEpro software [25]. This method evaluates evidence across four domains: risk of bias, inconsistency, indirectness, imprecision, and publication bias, rating each outcome as high, moderate, low, or very low certainty [26].

### Statistical analysis

Meta-analyses were performed for dichotomous outcomes using pooled odds ratios (ORs) with 95% confidence intervals (CIs). A Fixed or random-effects model (DerSimonian–Laird) was applied when substantial heterogeneity was present (I² >50%); otherwise, a fixed-effects model was used. Statistical heterogeneity was quantified using the I² statistic [27]. Publication bias was assessed through visual inspection of funnel plots and Egger’s regression test [28]. Pre-specified subgroup analyses compared EVT + IVT versus EVT alone. All statistical analyses were performed using Review Manager (RevMan) version 5.4 software.

## Results

### Study Selection

The systematic literature search identified 686 records from all the electronic databases. After removal of 95 duplicate citations, 591 unique records were screened based on titles and abstracts. Of these, 536 records were excluded for irrelevance, nonrandomized design, inappropriate comparators, or non–acute ischemic stroke populations. The remaining 55 articles underwent full-text review to assess eligibility. After full-text assessment, 44 studies were excluded due to non-randomized study designs, lack of relevant outcome data, overlapping populations, or failure to meet predefined inclusion criteria. Finally, 11 RCTs [29–39] enrolling a total of 4,419 participants satisfied all the eligibility criteria and were included in the final quantitative synthesis. The study selection process and reasons for exclusion at each stage are detailed in the PRISMA flow diagram (**Figure 1**) and PRISMA checklist is provided in **Supplementary Table 2**.

**Figure 1.**
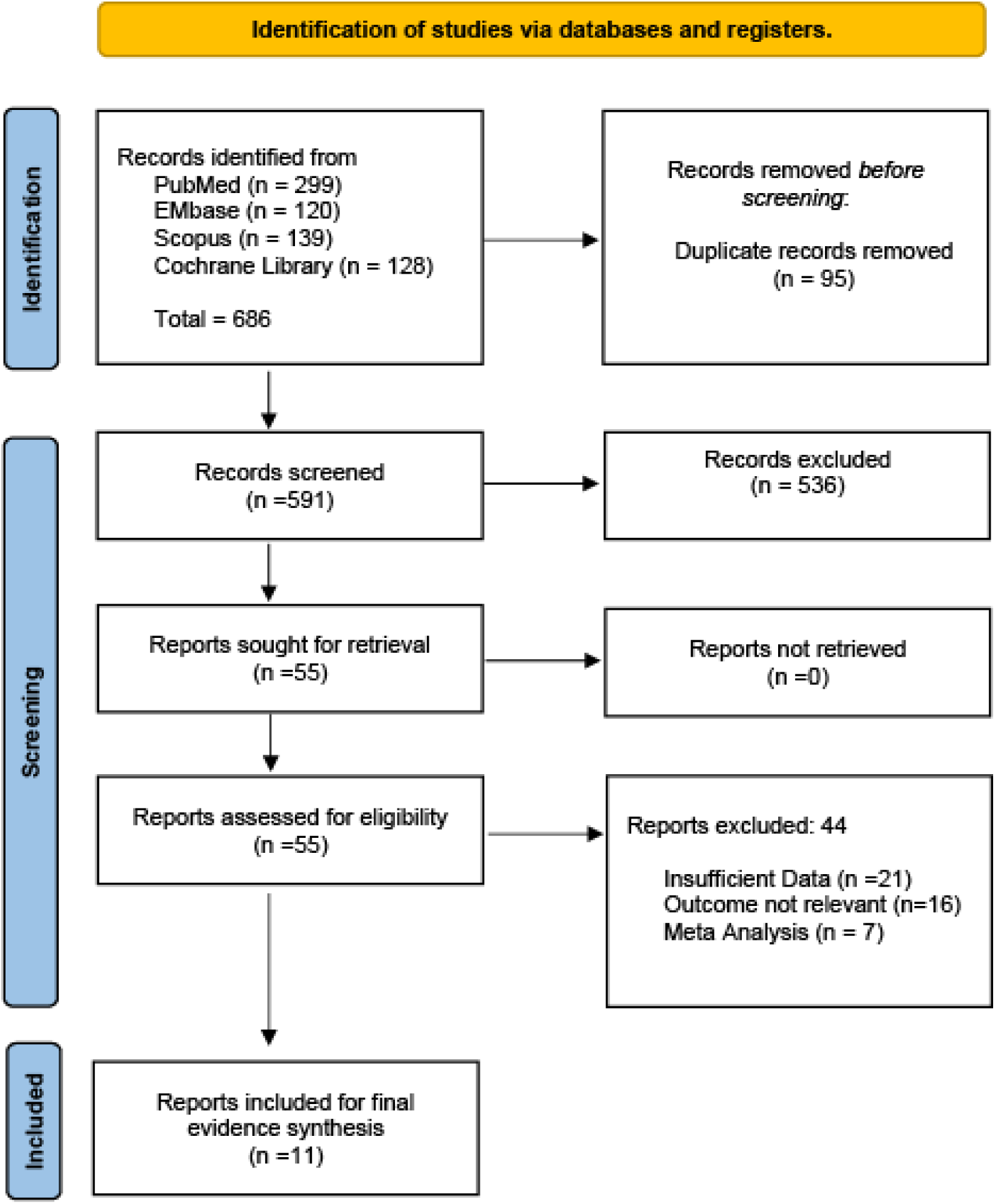
PRISMA flow diagram illustrating the study selection process

### Study Characteristics

Eleven multicentric RCTs [29–39] published between 2015 and 2026 were included, enrolling a total of 4,419 patients. The trials were conducted across North America, Europe, Asia, and Australia, with individual study periods ranging from 2010 to 2024. Enrolment windows varied from within 4.5 hours to 24 hours after symptom onset, with the majority of studies recruiting patients within 6 hours. Most trials focused on large-vessel occlusions in the anterior circulation, primarily involving the internal carotid artery and middle cerebral artery, whereas a limited number included basilar artery occlusions. Sample sizes per treatment arm ranged from 101 to 377 participants, with patients allocated to bridging therapy (EVT + IVT) vs. EVT alone.

Overall, participants were predominantly older adults, with median or mean ages ranging from approximately 65 to 76 years. Baseline stroke severity was comparable between groups, reflecting moderate-to-severe ischemic stroke, with NIHSS scores generally spanning from 10 to 22 at presentation. Where reported, baseline ASPECTS values typically ranged between 7 and 10, indicating limited early ischemic changes and relative preservation of salvageable brain tissue. Across trials, successful recanalization was uniformly assessed using standardized angiographic criteria, most commonly defined as achieving a TICI/mTICI/eTICI grade of 2b–3. sICH was defined using established and trial-specific criteria, including ECASS II/III, NINDS, Heidelberg Bleeding Classification, or a neurological deterioration threshold (typically an increase of ≥4 points on the NIHSS). The intervention arms comprised bridging therapy with intravenous thrombolysis—using alteplase (0.25–0.9 mg/kg) or tenecteplase—followed by EVT employing contemporary stent retrievers and/or aspiration techniques, including Solitaire and Trevo systems, while comparator arms received EVT alone. Functional recovery, rates of successful recanalization, all-cause mortality at 90 days, and symptomatic intracranial hemorrhage were the most consistently reported outcomes across trials. Detailed study-level baseline characteristics, including design, population, interventions, and outcome definitions, are summarized in **Table 1**.

**Table 1.** Baseline Characteristics of included RCTs comparing bridging therapy with EVT alone, including study design, sample size, and key outcomes.

| S. No. | Author (Year) | Country | Study Period | Enrollment Window (from symptom onset) | Target Vessel / Occlusion Site | Intervention Group | Device/tpA type dose | Sample Size (n) | Age (Mean ± SD or Median [IQR]) | Sex (M/F) | Baseline NIHSS (Mean ± SD or Median [IQR]) | ASPECT S | Recanalization Criteria (TICI/TI MI/mTICI I) | sICH Definition | Outcome Measured |
| --- | --- | --- | --- | --- | --- | --- | --- | --- | --- | --- | --- | --- | --- | --- | --- |
| 1. | Goyal *et al.* 2015[38] | Canada | Feb 2013 - Oct 2014 | within 12 hrs. | ICA, MCA | EVT+ IVT | Alteplase (0.9 mg/kg) | 165 | 71 (60-81) | 79/86 | 16 (13-20) | 9 (8-10) | TICI (2b-3) | NIHSS≥ 4 | FR, SRR, mortality & sICH |
|  |  |  |  |  |  | EVT | Solitaire™ stent retriever | 150 | 70 (60-81) | 71/79 | 17 (12-20) | 9 (8-10) |  |  |  |
| 2. | Gariel *et al.* 2018[29] | France | Oct 2015 – Oct 2016 | within 6 hrs | ICA, MCA (M1, M2) | EVT+ IVT | Alteplase (0.9 mg/kg) | 250 | 68.7±14.8 | 142/108 | 17 (12.5-20) | 7 (6-9) | mTICI (2b-3) | ECASS III | FR, SRR, mortality & sICH |
| | | | | | | EVT | Stent retrievers / aspiration | 131 | 72.2±13.0 | 65/66 | 18 (11-21) | 7 (5-9) | | | |
| 3. | Yang *et al.* 2020[30] | China | July 2019 – Sept 2019 | within 4.5 hrs | ICA, MCA (M1, M2) | EVT+IVT | Alteplase (0.9 mg/kg) | 329 | 69 (61-76) | 181/148 | 17 (14-22) | 9 (7-10) | eTICI (2b-3) | HBC | FR, SRR, mortality & sICH |
|  |  |  |  |  |  | EVT | Stent retriever | 327 | 69 (61-76) | 189/138 | 17 (12-21) | 9 (7-10) |  |  |  |
| 4. | Langezaal *et al.* 2021[31] | Netherland | Oct 2011 - Dec 2019 | within 6 hrs. | BA | EVT+IVT | Alteplase (0.9 mg/kg) | 146 | 67.2±11.9 | 96/150 | 22.1 | NR | mTICI (2b-3) | HBC | FR, mortality & sICH |
| | | | | | | EVT | Stent retrievers / aspiration | 154 | 66.8±13.1 | 100/54 | 21.9 | NR | | | |
| 5. | LeCouffe *et al.* 2021[39] | Netherland | Oct 2020 – Feb 2021 | within 4.5 hrs | ICA, MCA (M1, Proximal M2) | EVT+ IVT | Alteplase (0.9 mg/kg) | 266 | 69 (61-77) | 144/122 | 16 (10-20) | 9 (8-10) | eTICI (2b-3) | HBC | FR, SRR, mortality & sICH |
|  |  |  |  |  |  | EVT | Stent retriever | 273 | 72 (62-80) | 161/112 | 16 (10-20) | 9 (8-10) |  |  |  |
| 6. | Suzuki *et al.* 2021[36] | Japan | Jan 2017 – Oct 2019 | within 4.5 hrs | ICA, MCA (M1, M2) | EVT+ IVT | Alteplase (0.6 mg/kg) | 103 | 76 (67-80) | 72/31 | 17 (12-22) | 4 | TICI grade ≥2b | NINDS | FR, SRR, mortality & sICH |
|  |  |  |  |  |  | EVT | Stent retrievers / aspiration | 101 | 74 (67-80) | 56/45 | 19 (13-23) | 4 |  |  |  |
| 7. | Zi *et al.* 2021[32] | China | May 2018 – May 2020 | within 4.5 hrs | ICA, MCA (M1, M2) | EVT+ IVT | Alteplase (0.9 mg/kg) | 118 | 70 (60-78) | 66/52 | 16 (13-20) | 117 | mTICI (2b-3) | ECASS II | FR, SRR, mortality & sICH |
|  |  |  |  |  |  | EVT | Stent retrievers, thromboaspiration | 116 | 70 (60-77) | 66/50 | 16 (12-20) | 115 |  |  |  |
| 8. | Fischer *et al.* 2022[33] | Europe | Nov 2017 – May 2021 | within 4.5 hrs | ICA, MCA (M1, M2) | EVT+ IVT | Alteplase (0.9 mg/kg) | 207 | 72 (65-81) | 103/104 | 17 (12-20) | 8 (7-9) | mTICI (2b-3) | HBC | FR, SRR, mortality & sICH |
|  |  |  |  |  |  | EVT | Solitaire | 201 | 73 (64-81) | 96/105 | 17 (13-20) | 8 (7-9) |  |  |  |
| 9. | Mitchell *et al.* 2022[34] | Australia | Jun 2018 – Jul 2021 | within 4.5 hrs | ICA, MCA (M1, M2), BA, TE | EVT+ IVT | Alteplase/tenecteplase (0.9 mg/kg) | 147 | 69 (60-79) | 88/59 | 15 (10-20) | 10 (9-10) | mTICI (2b-3) | NIHSS≥ 4 | FR, SRR, mortality & sICH |
|  |  |  |  |  |  | EVT | Trevo | 146 | 70 (61-78) | 78/68 | 15 (11-20) | 10 (9-10) |  |  |  |
| 10. | Doheim *et al.* 2025[35] | USA | Mar 2014 – Dec 2018 | within 24 hrs. | ICA, MCA (M1, M2) | EVT+ IVT | Alteplase/Tenecteplase (0.9 mg/kg) | 377 | 71 (61-79) | 208/169 | 10 (7-16) | 10 (8-10) | mTICI (2b-3) | HBC | FR, SRR, mortality & sICH |
|  |  |  |  |  |  | EVT | Trevo stent-retriever (Stryker) | 162 | 74 (65-81) | 84/78 | 11 (6-18) | 10 (9-10) |  |  |  |
| 11. | Qiu *et al.* 2025[37] | China | May 2022– Sept 2024 | within 4.5 hrs | ICA, MCA (M1, M2), VBA | EVT+ IVT | Tenecteplase (0.25 mg/kg) | 278 | 70 (60–77) | 161/117 | 16 (12–20) | 8 (6–9) | eTICI (2b-3) | HBC | FR, SRR, mortality & sICH |
|  |  |  |  |  |  | EVT | Stent retrievers/thromboaspiration/balloon angioplasty | 272 | 70 (63–76) | 159/113 | 16 (12–20) | 8 (6–9) |  |  |  |

**Abbreviations:** AIS – Acute Ischemic Stroke; ST – Standard Treatment ; SR-Stent Retriever; Asp=Aspiration; PH – Parenchymal Hemorrhage; IAS – Intra-arterial Stroke; LVO – Large Vessel Occlusion; MCA – Middle Cerebral Artery; ICA – Internal Carotid Artery; ICO – Intracranial Occlusion; IAO – Intracranial Arterial Occlusion; BAO – Basilar Artery Occlusion; LAO – Large Artery Occlusion; VBA – Vertebro-Basilar Artery; PCA – Posterior Cerebral Artery; hrs. – Hours; CT – Computerized Tomography; MRA – Magnetic Resonance Angiography; NIHSS – National Institutes of Health Stroke Scale; SAE – Serious Adverse Event; FR – Functional Recovery; SRR – Successful Recanalization Rate; sICH – Symptomatic Intracranial Hemorrhage; mTICI – Modified Thrombolysis in Cerebral Infarction; TICI – Thrombolysis in Cerebral Infarction; TIMI – Thrombolysis in Myocardial Infarction; FPE – First Pass Effect; mAOL – Modified Arterial Occlusive Lesion; ECASS – European Cooperative Acute Stroke Study; NINDS – National Institute of Neurological Disorders and Stroke; SITS-MOST – Safe Implementation of Thrombolysis in Stroke-Monitoring Study; HBC – Heidelberg Bleeding Classification .

## Main Results

### Good functional recovery (mRS 0–2 at 90 days)

EVT + IVT demonstrated a trend toward improved functional independence at 90 days compared with EVT alone (OR=1.25, 95% CI; 1.02 to 1.53), with **substantial heterogeneity** across studies (I² = 62%) (***Figure 2a***).

**Figure 2.**
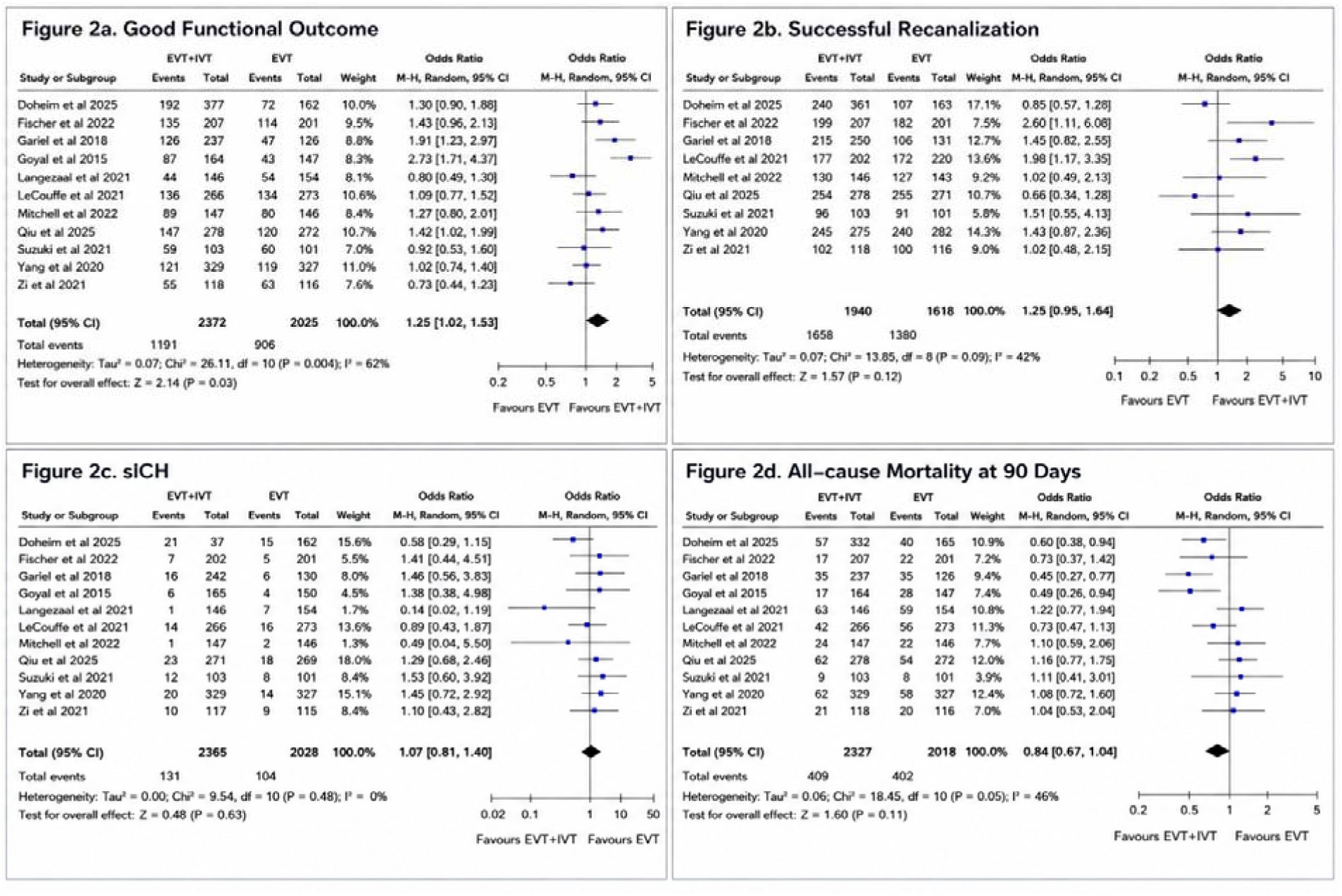
(a-d). Forest plot showing the pooled effect estimates for (a) Good functional recovery (mRS 0–2 at 90 days); (b) successful recanalization (mTICI 2b–3); (c) risk of symptomatic intracranial hemorrhage (sICH), and (d) all-cause mortality at 90 days in patients receiving bridging therapy versus EVT alone.

### Successful recanalization (mTICI 2b–3)

Bridging therapy was associated with significantly higher odds of successful recanalization than EVT alone (OR=1.25, 95% CI; 0.95 to 1.64), with moderate heterogeneity (I² = 42%) (***Figure 2b***).

### Symptomatic intracranial hemorrhage (sICH)

There was no significant difference in the risk of sICH between EVT + IVT and EVT alone (OR=1.07, 95% CI; 0.81 to1.40), with no heterogeneity (I² = 0%) (***Figure 2c***).

### All-cause mortality at 90 days

EVT + IVT was associated with lower 90-day all-cause mortality, although the difference did not reach statistical significance (OR=0.84, 95% CI; 0.67 to 1.04), with moderate heterogeneity (I² = 46%) (***Figure 2d***).

### Publication Bias

No evidence of publication bias was observed for any outcome. Funnel plots were visually symmetrical, and Egger’s tests were non-significant for functional independence at 90 days (p=0.98), successful recanalization (p=0.96), sICH (p=0.36), and all-cause mortality at 90 days (p=0.55), supporting the robustness of the pooled estimates (**Figure 3a–d**).

**Figure 3.**
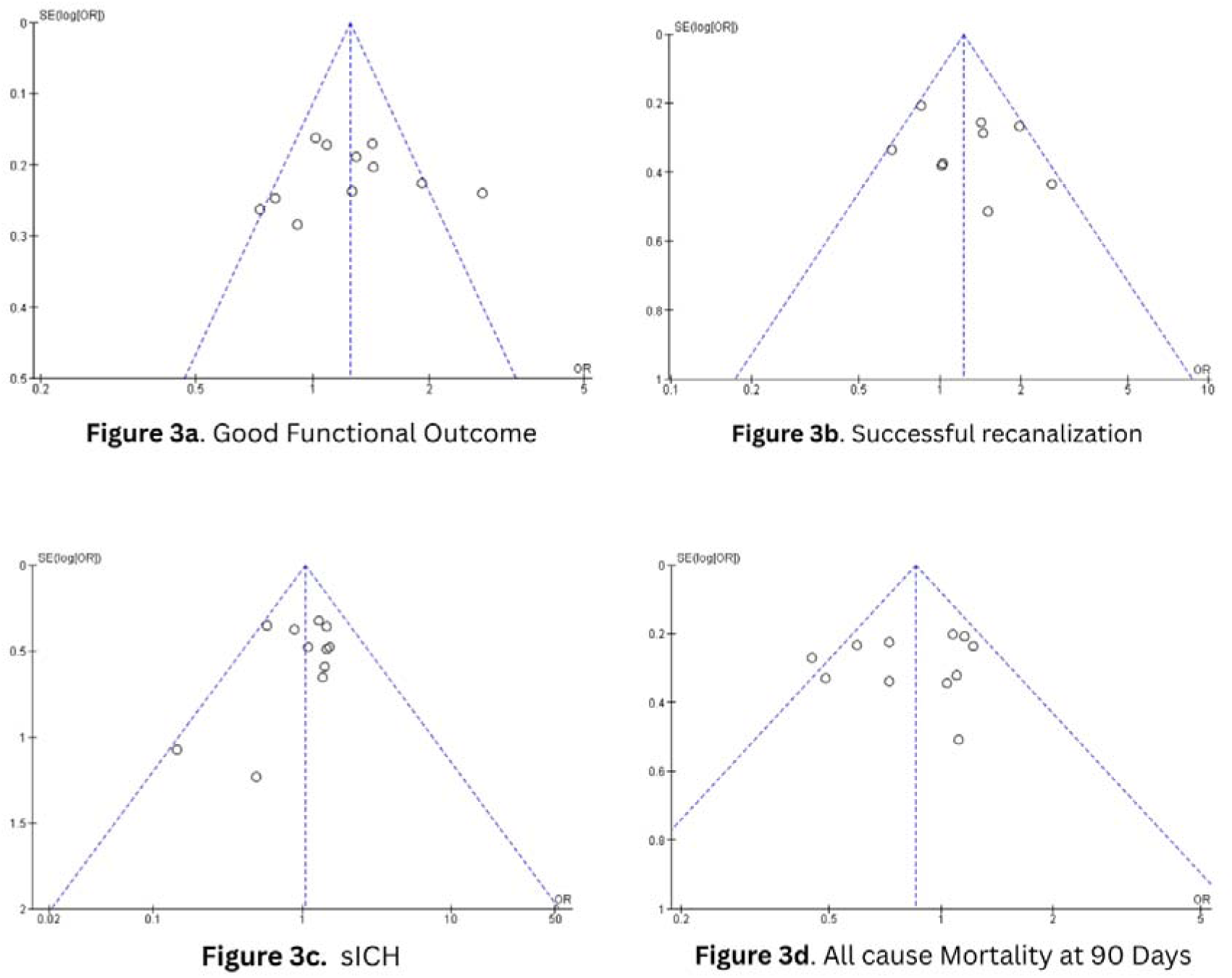
(a-d). Funnel plot assessing publication bias (a) Good functional recovery (mRS 0–2 at 90 days); (b) successful recanalization (mTICI 2b–3); (c) risk of symptomatic intracranial hemorrhage (sICH) and (d) all-cause mortality at 90 days in patients receiving bridging therapy versus EVT alone.

### Risk of Bias

The overall risk of bias assessment across the included studies demonstrated a predominantly low risk of bias in most methodological domains (**Figure 4a–b**). All studies were judged to have a low risk of bias for random sequence generation, allocation concealment, blinding of outcome assessment, incomplete outcome data, and selective reporting.

**Figure 4.**
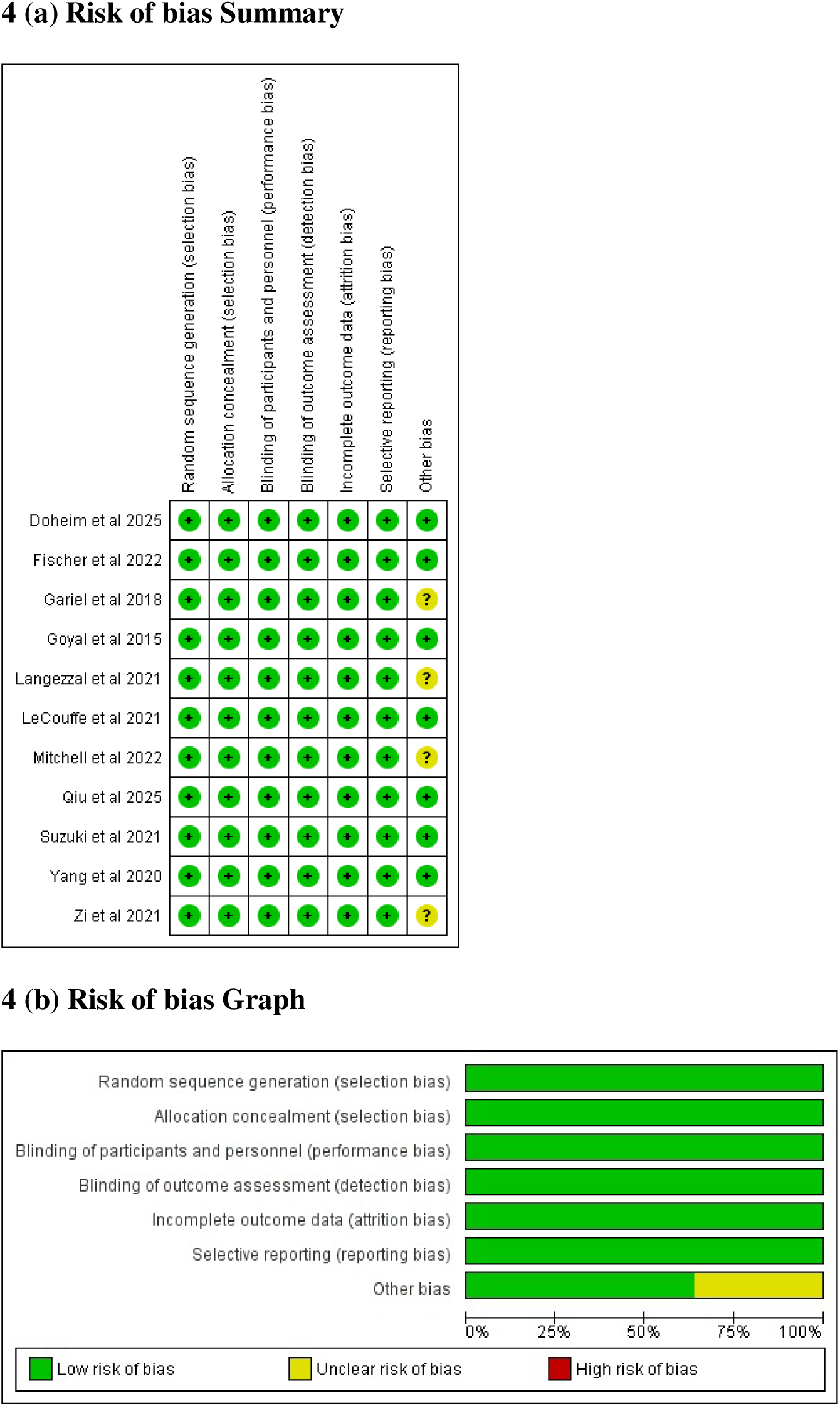
(a-b). Risk of bias assessment (a) Risk of bias Summary and (b) ROB graph for each included study based on the Cochrane Risk of Bias 2.0 tool.

For blinding of participants and personnel (performance bias), most studies were rated as having low risk of bias, as the intervention administration inherently required treating clinicians to be aware of the assigned treatment, making complete blinding challenging. Nevertheless, appropriate measures were implemented to minimize potential bias. Similarly, the domain assessing other sources of bias demonstrated a mixed pattern, with approximately half of the studies rated as low risk and the remaining studies assessed as having unclear risk of bias.

### Certainty of Evidence

GRADE assessment demonstrated moderate certainty of evidence for good functional recovery and successful recanalization with bridging therapy compared with EVT alone, with downgrading driven primarily by between-study heterogeneity (**Figure 5a–b**). The certainty of evidence for sICH was high, reflecting consistent effect estimates and no signal of increased bleeding risk (**Figure 5c**). Evidence for all-cause mortality at 90 days was of moderate certainty, downgraded due to imprecision and heterogeneity, although the pooled estimate favoured bridging therapy (**Figure 5d**).

**Figure 5.**
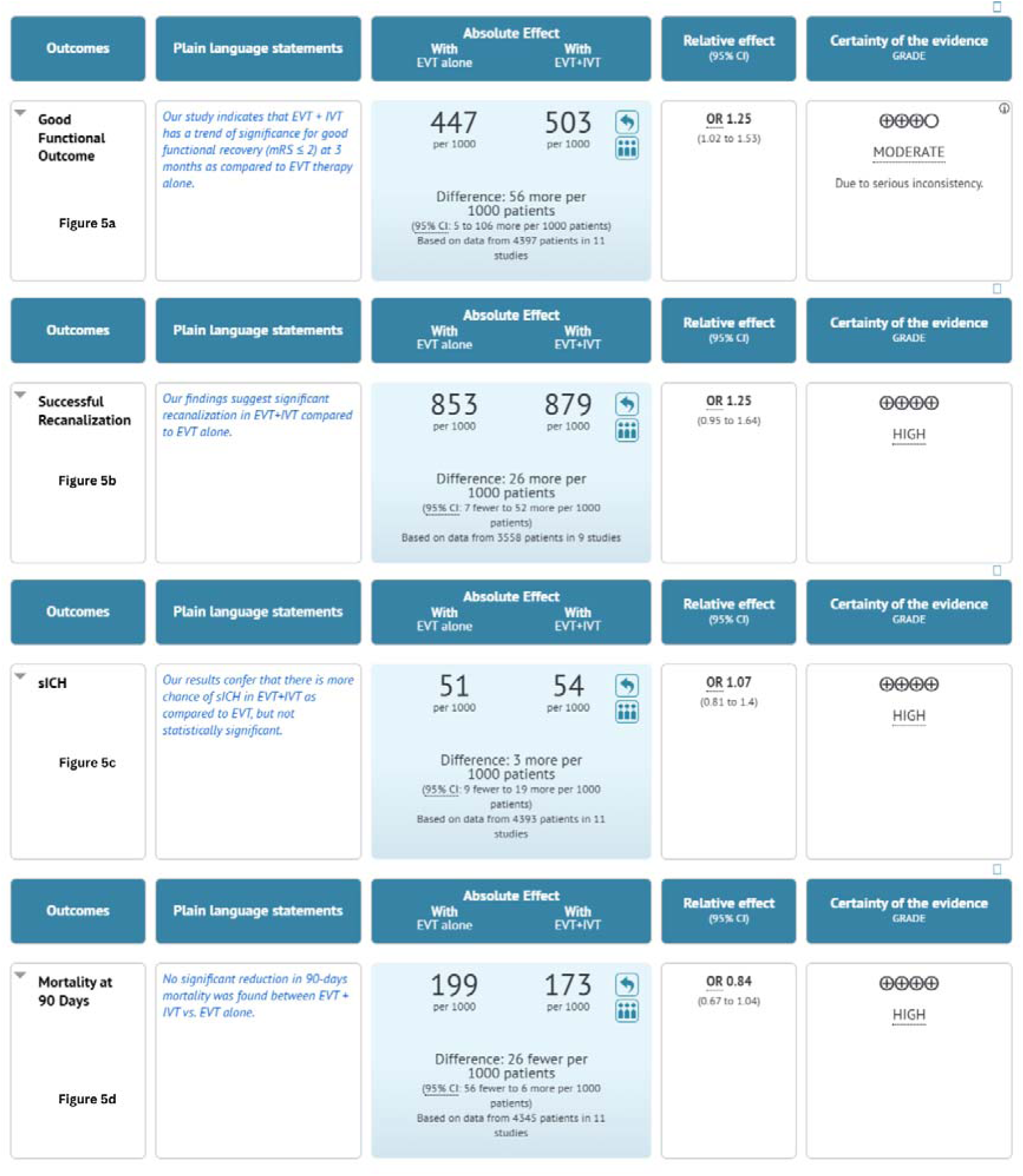
(a-d). GRADE summary of findings table presenting the quality of evidence and pooled effect estimates for (a) Good functional recovery (mRS 0–2 at 90 days); (b) successful recanalization (mTICI 2b–3); (c) risk of symptomatic intracranial hemorrhage (sICH) and (d) all- cause mortality at 90 days in patients receiving bridging therapy versus EVT alone.

### Discussion

In this comprehensive systematic review and meta-analysis of 11 RCTs involving 4,419 patients with acute ischemic stroke due to LVO, bridging therapy IVT before EVT was associated with significantly better functional independence at 90 days compared with EVT alone. In addition, bridging therapy demonstrated favourable trends toward improved recanalization and lower mortality without a significant increase in sICH. These findings support the continued clinical relevance of IVT in the modern thrombectomy era and reinforce the role of combined reperfusion therapy in appropriately selected patients.

The role of IVT prior to EVT has remained one of the most debated issues in contemporary stroke care. While EVT is established as the standard treatment for LVO stroke, uncertainty persists regarding whether prior thrombolysis provides additional benefit beyond mechanical clot retrieval. The present findings suggest that IVT continues to offer incremental advantages, particularly in terms of early reperfusion and downstream microvascular restoration.

The observed improvement in functional outcomes may be explained by several biological mechanisms. Early administration of IVT may initiate partial thrombus dissolution before mechanical intervention, reduce clot burden, and soften clot consistency, thereby facilitating thrombectomy. Furthermore, IVT may dissolve distal emboli and improve microcirculatory perfusion, which cannot be directly addressed through EVT alone. These effects may contribute to improved tissue-level reperfusion and greater salvage of the ischemic penumbra, ultimately translating into better neurological recovery.

Importantly, the present analysis demonstrated no statistically significant increase in sICH with bridging therapy. Although IVT exposure was associated with a numerically higher hemorrhagic risk, the overall safety profile remained acceptable across trials. This finding is clinically important because concerns regarding hemorrhagic complications remain one of the primary arguments favoring direct EVT strategies. Our results indicate that, when administered according to established eligibility criteria and standardized protocols, bridging therapy maintains a favorable balance between efficacy and safety.

### Comparison with Previous Literature

The findings of this study are consistent with previous contemporary evidence syntheses evaluating bridging therapy in acute ischemic stroke. Huang et al. [19] reported that bridging thrombolysis remained associated with favourable functional outcomes without substantially increasing haemorrhagic complications in patients undergoing EVT. Similarly, earlier meta-analyses[17,18,20,21] comparing EVT alone versus EVT plus IVT suggested that direct thrombectomy was not clearly superior to bridging therapy, although those analyses were limited by heterogeneity in trial designs, non-inferiority margins, imaging protocols, and thrombolytic strategies.

Compared with earlier reviews, the present study provides updated and methodologically rigorous evidence by exclusively including randomized controlled trials conducted in the modern thrombectomy era. The inclusion of contemporary multicentric trials enhances the clinical relevance and generalizability of the findings across diverse healthcare settings and stroke systems of care.

### Clinical Implications

The results of this meta-analysis have important implications for clinical practice and healthcare systems. In many regions, particularly in low- and middle-income countries, access to thrombectomy-capable centres remains limited and delays in EVT are common. In such settings, initiation of IVT before transfer for EVT may provide early reperfusion benefits during the hyper-acute phase of ischemia and potentially reduce infarct progression while awaiting definitive mechanical intervention.

Even in centres with optimized EVT workflows and rapid door-to-groin times, the observed improvement in functional outcomes supports the continued use of bridging therapy when no contraindications to IVT exist. These findings are also concordant with current international stroke guidelines recommending administration of IVT before EVT in eligible patients.

### Strengths of the Study

The present study has several important strengths. First, only randomized controlled trials were included, minimizing selection bias and enhancing the reliability of pooled estimates. Second, the review was conducted according to PRISMA 2020 and Cochrane recommendations with prospective PROSPERO registration, ensuring methodological transparency and reproducibility. Third, the inclusion of recent trials reflects contemporary stroke care practices, advanced imaging selection, and newer-generation thrombectomy devices. Finally, the use of GRADE methodology allowed structured assessment of the certainty of evidence across outcomes.

### Limitations

Several limitations should be acknowledged. First, the analysis was based on trial-level rather than individual patient-level data, limiting detailed subgroup analyses according to stroke severity, collateral status, clot location, thrombus characteristics, or onset-to-treatment time. Second, variations in thrombectomy techniques, imaging selection criteria, thrombolytic agents, and definitions of recanalization across studies may have contributed to heterogeneity. Third, the limited number of studies evaluating tenecteplase-based bridging therapy restricted assessment of its comparative effectiveness. Finally, differences in follow-up protocols and reporting standards among studies may have influenced the precision of pooled estimates.

### Conclusion

Bridging therapy before EVT may improve functional outcomes and recanalization without increasing the risk of sICH, supporting its use as a reasonable treatment strategy in eligible patients with acute ischemic stroke. The findings of this meta-analysis reinforce the continued role of intravenous thrombolysis in the modern thrombectomy era and support current guideline recommendations favouring combined reperfusion therapy when IVT can be administered without delaying EVT.

## Data Availability

All data produced in the present work are contained in the manuscript and associated supplementary materials.

## Declaration

### Author contributions

AJ & MN: Study screening, Data curation, Writing, MN & SM: quality assessment & validation, DV: Editing the original draft, PK: Conceptualization, Formal analysis, writing, reviewing & editing

### Ethics statement

Ethical approval and informed consent were not required for this systematic review and meta-analysis.

## Acknowledgement

None

## Supplementary Tables

Supplementary Table 1. A detailed search strategy was used for each electronic database (PubMed, EMbase, Scopus, Cochrane Library).

Supplementary Table 2. PRISMA Checklist

