## Supplemental Table 1 for "The Great Recanalization Debate in Acute Ischemic Stroke: Direct Thrombectomy versus Bridging Therapy—A Meta-analysis of Randomized Controlled Trials"

**Supplementary Table 1.** A detailed **search strategy** was used for each electronic database (PubMed, Embase, Scopus, Cochrane Library).

| Databases | Search Key Terms | Search Results |
| --- | --- | --- |
| PubMed | (("Stroke"[MeSH Terms] OR "Brain Ischemia"[MeSH Terms] OR "Ischemic Stroke"[All Fields] OR "Acute Ischemic Stroke"[All Fields] OR ("auton intell syst"[Journal] OR "artif intell surg"[Journal] OR "ais"[All Fields])) AND ("Thrombectomy"[MeSH Terms] OR "Endovascular Procedures"[MeSH Terms] OR "Mechanical Thrombectomy"[All Fields] OR "Endovascular Therapy"[All Fields] OR "EVT"[All Fields] OR "Endovascular Treatment"[All Fields]) AND ("Thrombolytic Therapy"[MeSH Terms] OR "thrombolysis"[All Fields] OR ("tissue plasminogen activator"[Supplementary Concept] OR "tissue plasminogen activator"[All Fields] OR "alteplase"[All Fields] OR "tissue plasminogen activator"[MeSH Terms] OR ("tissue"[All Fields] AND "plasminogen"[All Fields] AND "activator"[All Fields])) OR "tPA"[All Fields] OR "tissue plasminogen activator"[All Fields] OR "IVT"[All Fields] OR "intravenous thrombolysis"[All Fields]) AND ("randomized controlled trial"[Publication Type] OR "randomized"[Title/Abstract] OR "randomised"[Title/Abstract] OR "RCT"[Title/Abstract] OR "clinical trial"[Publication Type]) AND 2015/01/01:2025/10/31[Date - Publication]) AND (randomizedcontrolledtrial[Filter]) | 299 |
| EMBASE | ('ischemic stroke'/exp OR 'acute ischemic stroke' OR ais) AND ('endovascular therapy'/exp OR 'mechanical thrombectomy'/exp OR 'endovascular treatment' OR evt) AND ('thrombolytic therapy'/exp OR thrombolysis OR alteplase OR 'tissue plasminogen activator' OR ivt) AND ('randomized controlled trial'/exp OR randomized:ti,ab OR randomised:ti,ab OR rct:ti,ab) AND [2015-2025]/py AND 'clinical trial'/it | 120 |
| Scopus | TITLE-ABS-KEY(("acute ischemic stroke" OR "ischaemic stroke" OR AIS OR "large vessel occlusion" OR LVO)AND("mechanical thrombectomy" OR "endovascular thrombectomy" OR EVT OR "endovascular therapy" OR "endovascular treatment") AND ("intravenous thrombolysis" OR "IV thrombolysis" OR IVT OR thrombolysis OR alteplase OR "tissue plasminogen activator" OR tPA) AND("bridging therapy" OR "bridge therapy" OR "bridging thrombolysis" OR "direct EVT" OR "EVT alone" OR "EVT only") AND(randomized OR randomised OR "randomized controlled trial" OR RCT)  ) AND (PUBYEAR > 2014 AND PUBYEAR < 2026) | 139 |
| Cochrane Library | (("acute ischemic stroke" OR "ischaemic stroke" OR AIS OR "large vessel occlusion" OR LVO))  AND  (("mechanical thrombectomy" OR "endovascular thrombectomy" OR "endovascular treatment" OR "endovascular therapy" OR EVT))  AND  (("intravenous thrombolysis" OR "IV thrombolysis" OR "IVT" OR "thrombolytic therapy" OR thrombolysis OR alteplase OR "tissue plasminogen activator" OR tPA))  AND  (("with" NEAR/3 "without" thrombolysis) OR "bridging therapy" OR "bridging thrombolysis" OR "direct EVT")  AND (randomized OR randomised OR "controlled clinical trial" OR "randomized controlled trial" OR RCT) | 128 |

**Limits to**: Humans And RCTs; Search done from 01^st^ Jan 2015 to 30^th^ April 2026
